# Real-world effectiveness of NVX-CoV2373 and BNT162b2 mRNA COVID-19 vaccination in South Korea

**DOI:** 10.1101/2024.11.26.24317925

**Authors:** Eunseon Gwak, Seung-Ah Choe, Kyuwon Kim, Erdenetuya Bolormaa, Jonathan Fix, Muruga Vadivale, Matthew D. Rousculp, Young June Choe

**Author notes:** **Co-Correspondence**: Young June Choe, MD, PhD. Department of Pediatrics, Korea University College of Medicine, 73 Goryeodae-ro, Seongbuk-gu, Seoul 02841, Korea, and Matthew D. Rousculp, PhD, MPH, Novavax, Inc., Gaithersburg, MD 20878, USA. Equally contributed to the manuscript (Eunseon Gwak, Seung-Ah Choe, Matthew D. Rousculp, Young June Choe).

## Abstract

**Objectives:** In February 2022, NVX-CoV2373 became available in South Korea; real-world effectiveness of multiple doses compared with mRNA-based vaccines has not been thoroughly evaluated.

**Methods:** This retrospective study identified NVX-CoV2373 and BNT162b2 recipients aged ≥12 years from the K-COV-N database. Vaccine groups were propensity score–matched based on demographic characteristics, Seoul capital area residence, income level, comorbidity/disability, prior SARS-CoV-2 infection, and prior vaccination dose/timing. Outcomes were any and severe (intensive-care-unit admission or death within 8 weeks of infection) laboratory-confirmed SARS-CoV-2 infection assessed from 7 days after the third and fourth dose. Adjusted hazard ratios (aHRs) from matched groups measured vaccine effectiveness up to a 180-day risk window.

**Results:** From February–December 2022, 923,833 NVX-CoV2373 and 1,286,604 BNT162b2 doses were administered. The 180-day risk-window aHRs (95% CI) for NVX-CoV2373 compared with BNT162b2 for any SARS-CoV-2 infection were 0.78 (0.76–0.79) post third dose and 0.86 (0.86–0.87) post fourth dose. The 180-day aHRs (95% CI) for severe infection were 0.73 (0.53–1.00) after the third dose and 1.21 (1.03–1.42) after the fourth dose.

**Conclusions:** NVX-CoV2373 demonstrated favorable and similar effectiveness against any and severe SARS-CoV-2 infection, respectively, compared with BNT162b2, with evidence of enhanced NVX-CoV2373 durability.

## Introduction

The continual emergence of new SARS-CoV-2 variants has resulted in a persistent global burden of infection and cases of COVID-19.(1) Several types of vaccines against SARS-CoV-2 have been developed, many of which are now authorized or approved by local regulatory agencies in Korea(2) and across the globe.(3) In 2022, authorized or approved SARS-CoV-2 vaccines included messenger ribonucleic acid (mRNA)–based BNT162b2 (Comirnaty^®^, Pfizer-BioNTech; approved) and mRNA-1273 (Spikevax™, Moderna, Inc.; authorized), adenoviral vector-based Ad26.COV2.S (Johnson & Johnson/Janssen; no longer available in the United States [US] as of June 2023), and protein nanoparticle NVX-CoV2373 (Nuvaxovid™, Novavax, Inc.; authorized).(3) The primary series of authorized mRNA and protein-based COVID-19 vaccines consist of two doses administered 28 or 21 days apart.(4–7) The Korean Disease Control and Prevention Agency (KDCA) and World Health Organization (WHO) recommend additional doses of COVID-19 vaccines for high-risk populations, including individuals aged >65 years (KDCA) or >50 years (WHO) and individuals who are immunocompromised.(8, 9) The immune-evading capabilities of emerging variants(10) has required updates to vaccine strain compositions,(11) including a bivalent (ancestral and variant, combined) vaccine in 2022 and a change to monovalent Omicron XBB.1.5–based vaccines for 2023–2024(12). The most recent guidance is to target the JN.1 lineage of SARS-CoV-2 for 2024–2025.(13)

Pivotal studies with NVX-CoV2373 demonstrated high efficacy against SARS-CoV-2 infection in the US, Mexico, and the United Kingdom.(14, 15) As NVX-CoV2373 has been globally integrated into national immunization programs, including in South Korea, it is imperative to assess its real-world effectiveness. Although randomized-controlled trials investigating NVX-CoV2373 or mRNA-based vaccines have separately reported vaccine efficacy,(14, 16, 17) cross-trial comparisons present confounding factors (eg, study design, patient population, timing of studies), preventing additional conclusions from being made. Furthermore, real-world data on the relative effectiveness of multiple doses of NVX-CoV2373 in the general population are limited to the primary series and/or smaller populations. (18–22)

In South Korea, studying vaccine effectiveness with data from 2022 provides a unique research opportunity. Beginning in February 2020, the South Korean government instituted active epidemiological investigations, strict isolation of affected individuals, and extensive public lockdowns, which led to the country experiencing minimal impact from the COVID-19 pandemic.(23) With the loosening of these restrictions in late 2021, South Korea quickly experienced the highest incidence rate (IR) of infection in the world. From mid-February to mid-April 2022, the SARS-CoV-2 infection cases exceeded 600,000/day, yet the country maintained one of the lower mortality rates globally. This relatively high infection rate persisted throughout most of 2022 and included varied waves of Omicron variants (BA.1/1.1, BA.2/2.3, BA.5/5.2).(24)

The primary goal of this retrospective cohort study was to estimate the relative effectiveness of third and fourth vaccine doses of NVX-CoV2373 compared with BNT162b2 in preventing any and severe laboratory-confirmed SARS-CoV-2 infection during the early Omicron variant dominance in South Korea(25) using population-based data.

## Methods

### Study design and population

This retrospective, observational study used the K-COV-N cohort,(26) which links the KDCA and National Health Insurance Service (NHIS) databases to provide COVID-19 vaccination and health outcome data. Anonymized data for vaccination date, dose, type, location, and lot number were linked to participant demographics and healthcare resource utilization (in respect to SARS-CoV-2 infection). The collected data were analyzed at the NHIS data analysis center.

This analysis included residents of Korea from February 1, 2022, through December 31, 2022, aged ≥12 years who received at least one dose of NVX-CoV2373 or BNT162b2. A minimum of 365 days of observation prior to the date of vaccination and complete medical outcome data were required. Heterologous or homologous booster (third or fourth) doses were permitted; however, primary series effectiveness was only assessed in individuals with homologous first and second doses.

The primary objective was evaluation of relative effectiveness of third and fourth doses of NVX-CoV2373 compared with matched individuals who received BNT162b2. The secondary objective was evaluation of effectiveness of a homologous primary series of NVX-CoV2373 compared with that of a matched group who received a homologous primary series of BNT162b2.

### Outcomes and Assessments

Outcomes of interest were any and severe laboratory-confirmed SARS-CoV-2 infection after vaccination. The primary outcome measure was laboratory-confirmed SARS-CoV-2 infection, determined through rapid antigen tests (RATs) or polymerase chain reaction (PCR) administered by healthcare professionals. Results from self-administered RATs were not reported to the government.(19) Hospitalization was not included as an outcome measure in the assessment of vaccine effectiveness due to South Korea public health policy during the study period, which mandated hospitalization of individuals testing positive for SARS-CoV-2 (for quarantine purposes) regardless of symptom severity.(27) Severe SARS-CoV-2 infection was defined as admission to an intensive care unit (ICU) or death due to and within 30 days of infection. ICU admissions attributed to SARS-CoV-2 infection/COVID-19 were identified using specific treatment codes for ICU admission, coupled with a primary or secondary diagnosis of SARS-CoV-2–related conditions (ICD-10 codes: B342, B972, Z208, Z290, Z115, U181, Z038, U071, U072, U08, U09, and U10). Given the unavailability of precise cause-of-death data, any death occurring within 8 weeks of a confirmed SARS-CoV-2 infection was assumed to be related to COVID-19. This 8-week window was established based on the upper range observed in previous studies for the time between symptom onset and COVID-19–related death.(28)

### Analyses

The date of vaccine dosing (third dose, fourth dose, or first dose of primary series) was considered the index date (Day 0). First, availability of third and fourth doses were March 7, 2022, and March 14, 2022, respectively, and any post-primary series doses received prior to these dates were considered invalid and subsequently excluded. Participant follow-up for vaccine effectiveness began 7 days after receipt of the third and fourth doses and 14 days after the second dose of the primary series.

### Statistical analysis

Individuals who received NVX-CoV2373 were matched to recipients of BNT162b2 through propensity scores, which were estimated based on demographic characteristics, relative level of income, residential region (Seoul capital area or not), disability registration, prior SARS-CoV-2 infection history, Charlson comorbidity index assessed based on the all diagnosis records from 2021 and month of COVID-19 vaccination. Income was categorized as medical aid beneficiary (no/lowest income) and then into quartiles of 10%–40%, >40%–60%, >60%–80%, and >80%–100%. Participants who received NVX-CoV2373 were matched to BNT162b2 recipients, with adjustments for confounding by balancing the baseline characteristics between the two groups using the absolute standardized difference (aSD), where a value >0.1 indicated a significant imbalance.

To assess relative vaccine effectiveness, IRs per 1000 person-days and adjusted hazard ratios (aHRs), for any and severe laboratory-confirmed infections, were calculated for the matched sets. Time-varying exposure to the vaccines was considered, following individuals from 7 days (14 days for primary series) post vaccination until COVID-19 diagnosis, death, end of follow-up (cumulative 30-days per risk window), receipt of an additional COVID-19 dose, or study end. Vaccine effectiveness was assessed every 30 days for up to 180 days of follow-up and in the following risk windows: 30, 60, 90, 120, 150, and 180 days. To quantify potential residual confounding in the vaccine effectiveness estimates, a negative control exposure (recent vaccination) was evaluated by calculating adjusted odds ratios (aORs) for any SARS-CoV-2 infection,(29) comparing NVX-CoV2373 recipients to matched BNT162b2 recipients during days 0–5 post vaccination; as there should not be an effect of either vaccine during this period, any observed association would indicate residual confounding at baseline. Statistical analyses were conducted using SAS^®^ 9.4.

### Ethical oversight

This study was conducted in compliance with ethical standards and relevant local regulations. Formal review by an institutional review board was not required for this type of research (Korea University Institutional Review Board exemption No. 2023AN0124).

## Results

### Population and doses administered

A total of 923,833 NVX-CoV2373 and 1,286,604 BNT162b2 doses administered to the national population of South Korea between February 1, 2022, and December 31, 2022, were included in this study. Among those vaccinated, 209,982 individuals were excluded from this analysis for not having received a homologous primary series (Fig. 1). After receipt of a homologous primary series, a heterologous or homologous third dose of NVX-CoV2373 was received by 127,275 individuals and BNT162b2 by 254,841 individuals; 505,607 and 978,874 individuals received a fourth dose of NVX-CoV2373 and BNT162b2, respectively. Propensity score matching (as described in the Methods) resulted in the following NVX-CoV2373 to BNT162b2 ratios for outcomes assessment: third dose, ∼1:2; fourth dose, 1:1; primary series, ∼3:1.

**Fig. 1.**
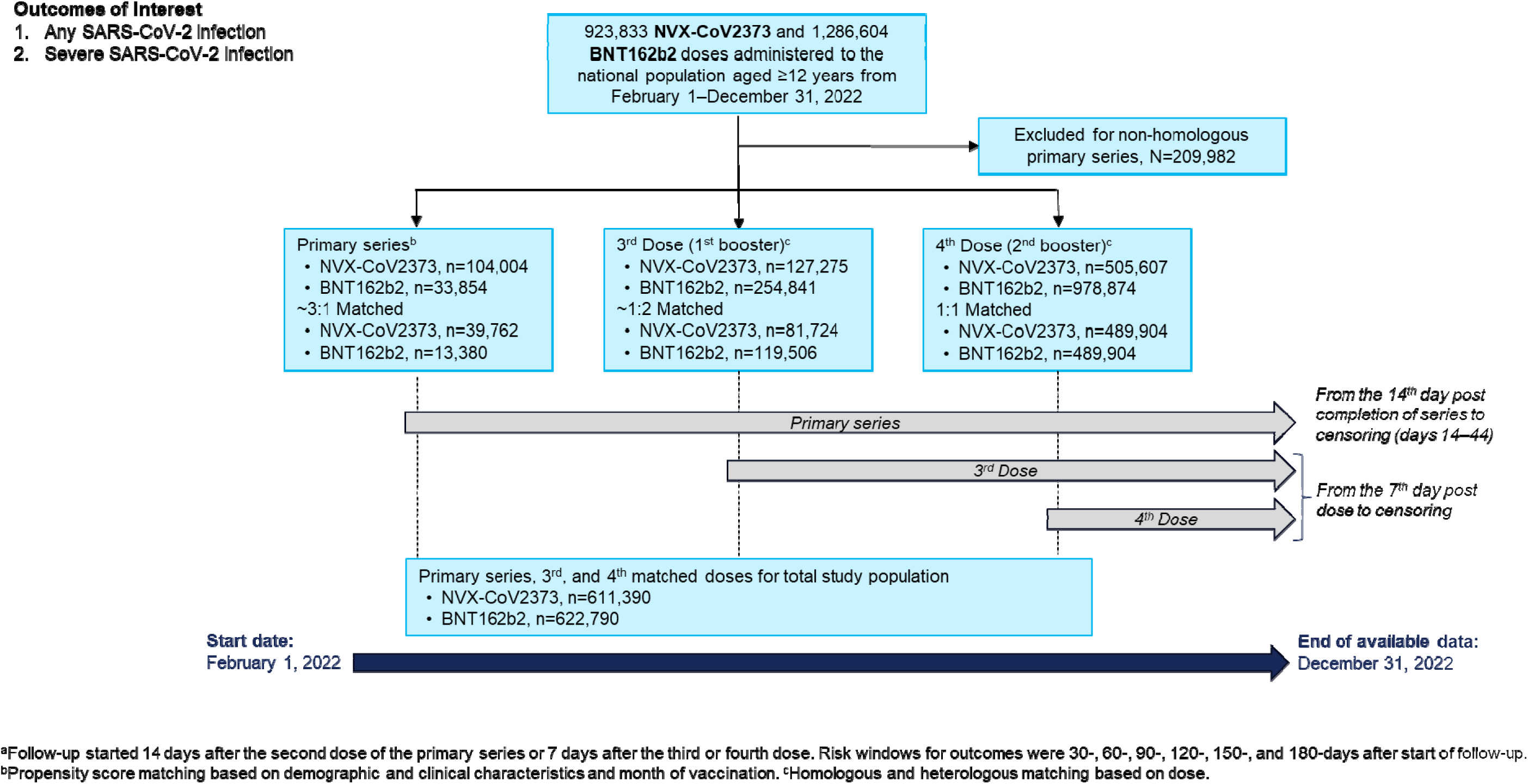
Study design for relative vaccine effectiveness comparing NVX-CoV2373 and BNT162b2.

There was a significantly greater proportion of matched third dose recipients of NVX-CoV2373 versus BNT162b2 who were aged 40–49 years, male, were employed, or were in the highest income quartile (Table 1). A significantly higher proportion of individuals who received a third dose of BNT162b2 than NVX-CoV2373 were aged 70–79 years or >80 years or were female. No significant differences were observed between the vaccine groups for the fourth-dose recipients (Table 1) or for the primary-series recipients (Supplement 1).

**Table 1.** Matched participant baseline characteristics for third and fourth doses, by vaccine type.

| Variable, n (%) | Third dose |  |  | Fourth dose |  |  |
| --- | --- | --- | --- | --- | --- | --- |
|  | NVX-CoV2373<br>(n=81,724) | BNT162b2<br>(n=119,506) | aSD<sup>a</sup> | NVX-CoV2373<br>(n=489,904) | BNT162b2<br>(489,904) | aSD<sup>a</sup> |
| Age |  |  |  |  |  |  |
| 12–17 | 20 (0.0) | 431 (0.4) | 0.05 | 0 (0.0) | 0 (0.0) | – |
| 18–24 | 9545 (11.7) | 15,834 (13.3) | 0.04 | 407 (0.1) | 521 (0.1) | 0.01 |
| 25–39 | 13,648 (16.7) | 24,257 (20.3) | 0.09 | 842 (0.2) | 1176 (0.2) | 0.01 |
| 40–49 | 22,359 (27.4) | 21,106 (17.7) | 0.24 | 3330 (0.7) | 2338 (0.5) | 0.02 |
| 50–59 | 18,044 (22.1) | 22,767 (19.1) | 0.08 | 84,240 (17.2) | 73,537 (15.0) | 0.07 |
| 60–69 | 11,966 (14.6) | 19,051 (15.9) | 0.04 | 213,277 (43.5) | 220,406 (45.0) | 0.03 |
| 70–79 | 3974 (4.9) | 9394 (7.9) | 0.13 | 132,076 (27.0) | 134,753 (27.5) | 0.01 |
| ≥80 | 2168 (2.7) | 6666 (5.6) | 0.15 | 55,732 (11.4) | 57,173 (11.7) | 0.01 |
| Sex |  |  |  |  |  |  |
| Male | 38,760 (47.4) | 49,730 (41.6) | 0.12 | 242,918 (49.6) | 243,113 (49.6) | <0.01 |
| Female | 42,964 (52.6) | 69,776 (58.4) | – | 246,986 (50.4) | 246,791 (50.4) | – |
| Non-capital area<sup>b</sup> | 39,655 (48.5) | 52,288 (43.8) | 0.10 | 285,394 (58.3) | 272,387 (55.6) | 0.05 |
| CCI ≥3 | 5045 (6.2) | 7371 (6.2) | <0.01 | 59,351 (12.1) | 60,001 (12.3) | <0.01 |
| Disabled | 4208 (5.2) | 7744 (6.5) | 0.06 | 58,714 (12.0) | 59,892 (12.2) | 0.01 |
| Employed | 38,719 (47.4) | 47,723 (39.9) | 0.15 | 138,183 (28.2) | 132,153 (27.0) | 0.03 |
| Income level<sup>c</sup> |  |  |  |  |  |  |
| Medical aid beneficiary | 2598 (3.2) | 5108 (4.4) | 0.06 | 26,921 (5.6) | 28,023 (5.8) | 0.01 |
| 1Q | 10,504 (13.1) | 14,722 (12.7) | 0.02 | 73,819 (15.3) | 74,188 (15.4) | <0.01 |
| 2Q | 12,945 (16.1) | 23,049 (19.8) | 0.09 | 93,727 (19.4) | 95,092 (19.7) | 0.01 |
| 3Q | 18,441 (23.0) | 28,875 (24.8) | 0.04 | 102,438 (21.2) | 103,770 (21.5) | 0.01 |
| 4Q | 35,772 (44.6) | 44,596 (38.3) | 0.13 | 186,389 (38.6) | 182,191 (37.7) | 0.02 |
| Obese (BMI >30kg/m<sup>2</sup>) | 2382 (2.9) | 2546 (2.1) | 0.05 | 11,797 (2.4) | 11,147 (2.3) | 0.01 |
| Current smoker | 5695 (7.0) | 7232 (6.1) | 0.04 | 27,631 (5.6) | 27,598 (5.6) | <0.01 |
| Prior SARS-CoV-2 infection | 12,078 (14.8) | 15,707 (13.1) | 0.05 | 114,233 (23.3) | 113,250 (23.1) | <0.01 |

<sup>a</sup>aSD values >0.1 indicate a significant imbalance between vaccine groups. <sup>b</sup>Indication of whether participants were living inside or outside of the Seoul capital area. <sup>c</sup>Income was categorized as medical aid beneficiary (no/lowest income) and then into quartiles of 10%–40%, >40%–60%, >60%–80%, and >80%–100%.
aSD, absolute standardized difference; BMI, body mass index; CCI, Charleson comorbidity index; Q, quartile.

### Incidence of SARS-CoV-2 infection after third or fourth COVID-19 vaccination

Within the 30-day risk window, SARS-CoV-2 infection occurred in 6455 (IR/1000 person-days=2.76, 95% CI: 2.70–2.83) participants who received a third dose of NVX-CoV2373 and in 9799 (2.86 [2.80–2.91]) participants who received a third dose of BNT162b2 (Table 2). There were 10 (IR=0.00 [95% CI: 0.00–0.01]) and 32 (0.01 [0.01–0.01]) individuals with severe SARS-CoV-2 infection in the 30-day risk window after the third dose of NVX-CoV2373 and BNT162b2, respectively. Of individuals who received a fourth COVID-19 vaccination, there were 11,306 (IR=0.78 [95% CI: 0.77–0.79]) NVX-CoV2373 recipients and 9115 (0.63 [0.61–0.64]) BNT162b2 recipients with any SARS-CoV-2 infection within the 30-day risk window; severe infection was observed in 30 (0.00 [0.00–0.00]) and 21 (0.00 [0.00–0.00]) recipients, respectively.

**Table 2.** Incidence rates (IRs) and adjusted hazard ratios (aHRs) for any and severe SARS-CoV-2 infection following third and fourth COVID-19 vaccine doses (presented by risk window since vaccination^a^).

| Risk window | Dose SARS-CoV-2 infection | NVX-CoV2373 |  |  | BNT162b2 |  |  | aHR (95% CI) |
| --- | --- | --- | --- | --- | --- | --- | --- | --- |
|  |  | Events, n | Time (1000 person-days) | IR (95% CI)<sup>b</sup> | Events, n | Time (1000 person-days) | IR (95% CI)<sup>b</sup> |  |
| 30 days | *Third* |  |  |  |  |  |  |  |
|  | Any | 6455 | 2337 | 2.76 (2.70–2.83) | 9799 | 3429 | 2.86 (2.80–2.91) | 0.94 (0.91–0.97) |
|  | Severe | 10 | 2451 | 0.00 (0.00–0.01) | 32 | 3585 | 0.01 (0.01–0.01) | 0.66 (0.32–1.35) |
|  | *Fourth* |  |  |  |  |  |  |  |
|  | Any | 11,306 | 14,504 | 0.78 (0.77–0.79) | 9115 | 14,551 | 0.63 (0.61–0.64) | 1.19 (1.16–1.23) |
|  | Severe | 30 | 14,696 | 0.00 (0.00–0.00) | 21 | 14,697 | 0.00 (0.00–0.00) | 1.37 (0.78–2.42) |
| 60 days | *Third* |  |  |  |  |  |  |  |
|  | Any | 9042 | 4545 | 1.99 (1.95–2.03) | 15,795 | 6615 | 2.39 (2.35–2.43) | 0.81 (0.79–0.83) |
|  | Severe | 16 | 4903 | 0.00 (0.00–0.00) | 68 | 7167 | 0.01 (0.01–0.01) | 0.51 (0.29–0.88) |
|  | *Fourth* |  |  |  |  |  |  |  |
|  | Any | 24,542 | 28,681 | 0.86 (0.85–0.87) | 23,621 | 28,789 | 0.82 (0.81–0.83) | 1.00 (0.98–1.02) |
|  | Severe | 64 | 29,391 | 0.00 (0.00–0.00) | 49 | 29,392 | 0.00 (0.00–0.00) | 1.34 (0.92–1.95) |
| 90 days | *Third* |  |  |  |  |  |  |  |
|  | Any | 11,318 | 6684 | 1.69 (1.66–1.72) | 20,046 | 9656 | 2.08 (2.05–2.11) | 0.79 (0.77–0.81) |
|  | Severe | 28 | 7353 | 0.00 (0.00–0.00) | 88 | 10,749 | 0.01 (0.01–0.01) | 0.69 (0.45–1.07) |
|  | *Fourth* |  |  |  |  |  |  |  |
|  | Any | 57,000 | 42,209 | 1.35 (1.34–1.36) | 62,023 | 42,274 | 1.47 (1.46–1.48) | 0.88 (0.87–0.89) |
|  | Severe | 135 | 44,081 | 0.00 (0.00–0.00) | 111 | 44,083 | 0.00 (0.00–0.00) | 1.27 (0.99–1.64) |
| 120 days | *Third* |  |  |  |  |  |  |  |
|  | Any | 14,495 | 8747 | 1.66 (1.63–1.68) | 25,253 | 12,560 | 2.01 (1.99–2.04) | 0.80 (0.78–0.81) |
|  | Severe | 37 | 9803 | 0.00 (0.00–0.00) | 115 | 14,329 | 0.01 (0.01–0.01) | 0.69 (0.48–1.01) |
|  | *Fourth* |  |  |  |  |  |  |  |
|  | Any | 91,114 | 54,631 | 1.67 (1.66–1.68) | 99,656 | 54,486 | 1.83 (1.82–1.84) | 0.86 (0.86–0.87) |
|  | Severe | 243 | 58,764 | 0.00 (0.00–0.00) | 202 | 58,768 | 0.00 (0.00–0.00) | 1.28 (1.06–1.55) |
| **150 days** | *Third* |  |  |  |  |  |  |  |
|  | Any | 19,677 | 10,683 | 1.84 (1.82–1.87) | 33,689 | 15,262 | 2.21 (2.18–2.23) | 0.78 (0.77–0.80) |
|  | Severe | 47 | 12,253 | 0.00 (0.00–0.00) | 140 | 17,908 | 0.01 (0.01–0.01) | 0.70 (0.50–0.97) |
|  | *Fourth* |  |  |  |  |  |  |  |
|  | Any | 102,982 | 66,366 | 1.55 (1.54–1.56) | 112,483 | 65,954 | 1.71 (1.70–1.72) | 0.86 (0.86–0.87) |
|  | Severe | 294 | 73,448 | 0.00 (0.00–0.00) | 261 | 73,453 | 0.00 (0.00–0.00) | 1.19 (1.01–1.41) |
| **180 days** | *Third* |  |  |  |  |  |  |  |
|  | Any | 23,820 | 12,467 | 1.91 (1.89–1.94) | 40,214 | 17,722 | 2.27 (2.25–2.29) | 0.78 (0.76–0.79) |
|  | Severe | 53 | 14,702 | 0.00 (0.00–0.00) | 148 | 21,488 | 0.01 (0.01–0.01) | 0.73 (0.53–1.00) |
|  | *Fourth* |  |  |  |  |  |  |  |
|  | Any | 106,377 | 77,899 | 1.37 (1.36–1.37) | 116,057 | 77,199 | 1.50 (1.49–1.51) | 0.86 (0.86–0.87) |
|  | Severe | 330 | 88,131 | 0.00 (0.00–0.00) | 289 | 88,138 | 0.01 (0.01–0.01) | 1.21 (1.03–1.42) |

<sup>a</sup>Risk windows began 7 days post dose and ran from days 7–37, 7–67, 7–97, 7–127, 7–157, and 7–187. <sup>b</sup>Calculated per 1000 person-days.

In the 180-day risk window after the third dose, any SARS-CoV-2 infection was detected in 23,820 (IR=1.91 [95% CI: 1.89–1.94]) recipients of NVX-CoV2373 and 40,214 (2.27 [2.25–2.29]) recipients of BNT162b2 (Table 2). The cumulative incidence of severe SARS-CoV-2 infection was 53 and 148 cases, respectively, for recipients of NVX-CoV2373 (IR=0.00 [95% CI: 0.00–0.00]) and BNT162b2 (0.01 [0.01–0.01]). Within the 180-day risk window for individuals who received a fourth dose of NVX-CoV2373, there were 106,377 (IR=1.37 [95% CI: 1.36–1.37]) individuals who experienced SARS-CoV-2 infection; 116,057 (1.50 [95% CI 1.49–1.51]) recipients of BNT162b2 had a reported infection. Cumulative cases of severe infection after the fourth dose were observed in 330 (IR=0.00 [95% CI: 0.00–0.00]) and 289 (0.01 [0.01–0.01]) recipients of NVX-CoV2373 and BNT162b2, respectively. Outcomes among each of the risk windows demonstrated a similar trend with one another (Table 2). Cumulative risk of any SARS-CoV-2 infection since the third and fourth dose of either vaccine is shown in Fig. 2.

**Fig. 2.**
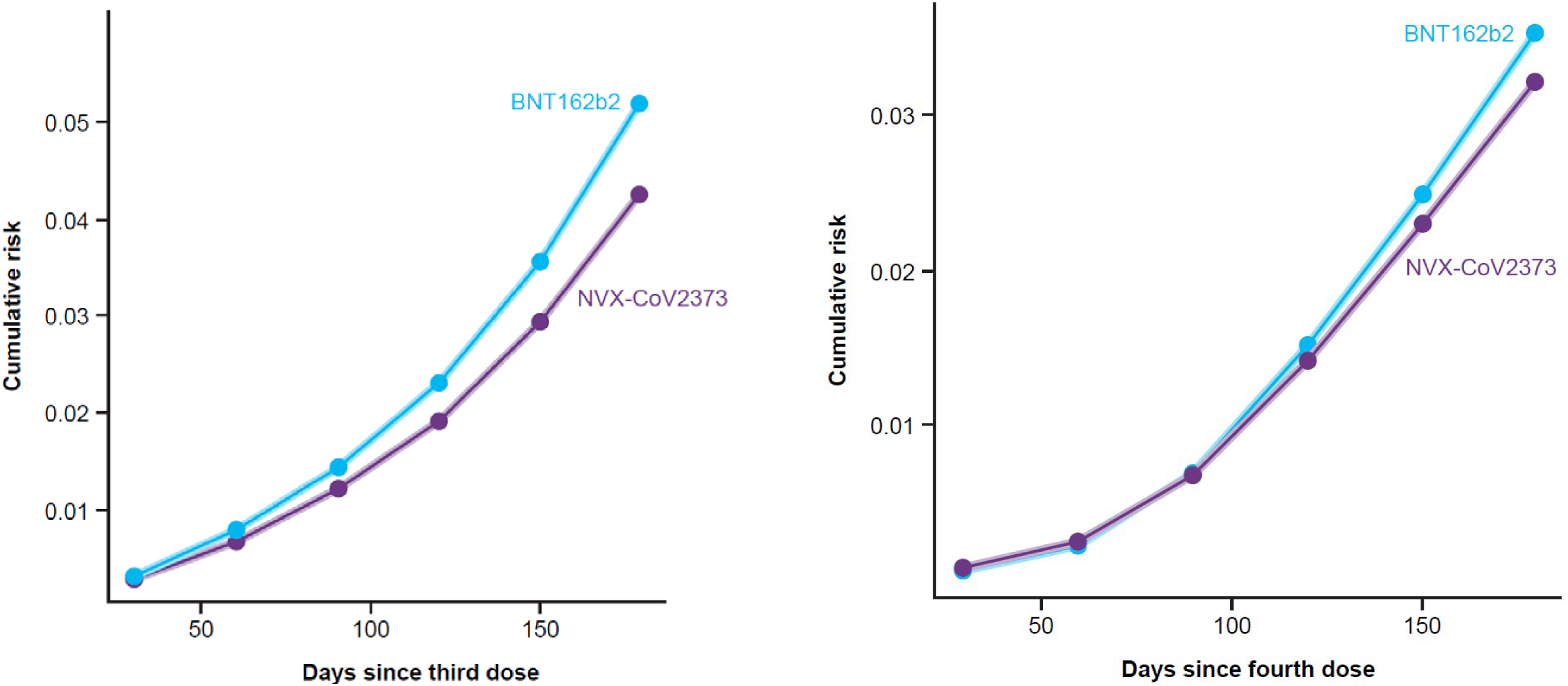
Cumulative risk of all SARS-CoV-2 infections after receipt of third dose (left) or fourth dose (right) of NVX-CoV2373 or BNT162b2. Shaded area presents 95% confidence intervals of incidences. Risk windows began 7 days post dose and ran from days 7–37, 7–67, 7–97, 7–127, 7–157, and 7–187.

### SARS-CoV-2 infection risk comparison among vaccination groups

The aHR (95% CI) for NVX-CoV2373 compared with BNT162b2 for any SARS-CoV-2 infection in the 30-day risk window after the third dose was 0.94 (95% CI: 0.91–0.97) and for severe infection was 0.66 (0.32–1.35; Table 2; Fig. 3). Risk estimates (aHR [95% CI]) observed for the 180-day risk window after the third dose were 0.78 (95% CI: 0.76–0.79) for any SARS-CoV-2 infection and 0.73 (0.53–1.00) for severe infection. Among individuals who received a fourth dose, the aHR of SARS-CoV-2 infection in the 30-day risk window was higher for recipients of NVX-CoV2373 compared with BNT162b2 (any infection=1.19 [95% CI: 1.16– 1.23]; severe infection=1.37 [0.78–2.42]). For the 180-day risk window after the fourth dose, NVX-CoV2373 compared with BNT162b2 was associated with a lower risk estimate for overall infection (aHR=0.86 [95% CI: 0.86–0.87]); for severe infection, there was an aHR of 1.21 (95% CI: 1.03–1.42). Incidence and hazard ratios of any and severe SARS-CoV-2 infections for homologous, primary series recipients of NVX-CoV2373 or BNT162b2 are available in Supplements 2 and 3. Cumulative rates are shown in Supplement 4.

**Fig. 3.**
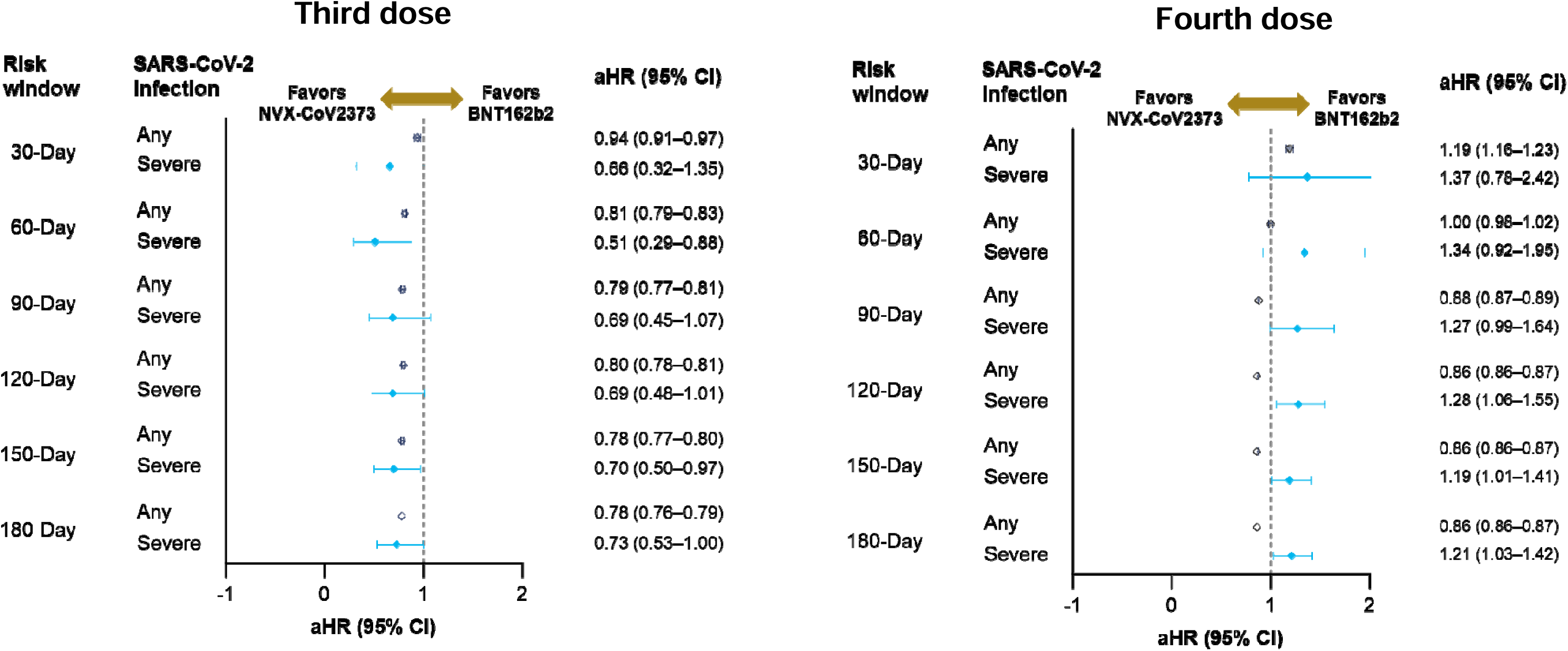
Adjusted hazard ratios (aHRs) of any or severe SARS-CoV-2 infection after a third or fourth dose of NVX-CoV-2373 or BNT162b2. Risk estimates after a third (left panel) or fourth (right panel) dose of NVX-CoV2373 versus BNT162b2 is displayed as aHR with 95% CI. Risk windows began 7 days post dose and ran from days 7–37, 7–67, 7–97, 7–127, 7–157, and 7–187. aHR values <1 favor protection from SARS-CoV-2 infection with NVX-CoV2373 and >1 favor BNT162b2.

### Evaluation of residual bias/confounding using the recently vaccinated period

The odds of any infection during days 0–5 post vaccination were generally similar between the NVX-CoV2373 and BNT162b2 cohorts for both the third dose and primary series (Supplement 5). For the fourth dose, the odds of any infection were significantly higher among the NVX-CoV2373 versus BNT162b2 group (aOR=1.16 [95% CI: 1.07–1.25]).

## Discussion

This analysis represents the largest real-world, population-based assessment of NVX-CoV2373 effectiveness to date. IRs of any laboratory-confirmed SARS-CoV-2 infection between the NVX-CoV2373 and BNT162b2 groups across the defined risk windows, after heterologous or homologous third and fourth doses, favored the effectiveness of the NVX-CoV2373 vaccine. No clinically meaningful difference was observed across the IRs for severe SARS-CoV-2 infection. Over the 180-day follow-up, the relative effectiveness of NVX-CoV2373, compared with BNT162b2, appeared to improve with time. The clinical meaning and public health implications of these findings require further investigation.

The observed improvements in vaccine effectiveness over time for NVX-CoV2373 compared with BNT162b2 may reflect the rapid waning of protective effects of the mRNA-based vaccines, which has been previously reported.(30) In the systematic review and meta-analysis by Menegale et al., vaccine effectiveness against symptomatic disease from 1 to 9 months post primary series decreased from 80% to 50% against Delta variant infection and from 53% to 9% against Omicron infection. Diminished vaccine effectiveness over time is a concern and long-term real-world monitoring will be valuable in determining durability of responses to initial (primary series) and subsequent vaccinations, as well as among the different formulations and vaccine updates. Evidence for enhanced durability of NVX-CoV2373 compared with mRNA vaccines has also been implied by the observed persistence of humoral responses over time. In an 8-month follow-up analysis of the COV-BOOST trial, a heterologous third dose of NVX-CoV2373 following a BNT162b2 primary series had a significantly slower decay rate of anti-spike immunoglobulin G levels compared with three homologous BNT162b2 doses.(31) While it is likely that serum antibody responses play a role in SARS-CoV-2 vaccine durability, there are other vaccine type–specific effector and memory responses that influence durability and cross-reactivity, which remain poorly understood and require additional research.

This study provides a unique perspective on outcomes assessment by vaccine history via the inclusion of heterologous doses after the primary series. While effectiveness outcomes reported in this study focused on booster doses, comparable levels of protection have been previously reported in observational investigations of vaccine-naive populations. A retrospective analysis accessing data in the K-COV-N database was conducted among adults without prior SARS-CoV-2 infection who received their first three doses of a COVID-19 vaccine from February–November 2022.(32) The 40-week assessment reported a low risk of any or severe SARS-CoV-2 infection with either NVX-CoV2373 or BNT162b2 (N≈3000 for each group). Another study reported low rates of COVID-19 among matched cohorts of Korean adults who received a homologous primary series of NVX-CoV2373 (N=47,078) or BNT162b2 (N=7561).(33) Data from February–April 2022 indicated similar rates of COVID-19 at 30, 60, and 90 days post primary series and a 22-week risk ratio for NVX-CoV2373 to BNT162b2 of 1.1. In a separate assessment of COVID-19 vaccine–naive adolescents (aged 12–17 years) in South Korea who received both doses of a homologous primary series (NVX-CoV2373 or BNT162b2) from September–December 2022, there was noninferiority among the two matched cohorts (each N=465) for SARS-CoV-2 infection.(19) Notably, the observation periods for each of these studies(19, 32, 33) were when early Omicron variants (eg, BA.1 BA.2, BA.2.3, and BA.4/5) were predominant,(25) suggesting benefits of the prototype vaccine against variant strains with spike mutations.

Propensity score matching was used to control for baseline differences between the vaccinated populations; however, residual and unmeasured confounding remained a potential concern. Residual confounding was evaluated through a negative control analysis, comparing the odds of SARS-CoV-2 infection during days 0–5 post vaccination between each group, at which point the vaccine would not yet be effective. While this approach does not address time-varying confounding or other biases (eg, selection bias), it helps demonstrate how well the control variables in the statistical model account for baseline differences between the vaccinated populations. The odds of infection were higher for NVX-CoV2373 than BNT162b2 recipients following the fourth dose, suggesting that fourth-dose NVX-CoV2373 recipients might have had a higher baseline risk for medically attended SARS-CoV-2 infection and/or could have been more likely to seek healthcare compared with their matched counterparts.

One limitation of this study is the timing of the study period, which coincided with the largest COVID-19 outbreak in South Korea (mid-February to mid-April 2022), driven by the Omicron variant, and resulting in an increase in cases to the highest incidence (>600,000 cases/day) observed at that time.(23) This extraordinary surge in infections placed immense strain on the healthcare system, potentially impacting the accuracy of data collection and reporting. Moreover, the widespread transmission during this period might have influenced individuals’ behaviors and adherence to preventive measures, further complicating the assessment of vaccine effectiveness; however, robust community testing and uniform testing recommendations in South Korea should have helped mitigate differences in health-seeking behavior among the populations included. Another limitation is that only prototype vaccine formulations targeting ancestral SARS-CoV-2 were evaluated. There was likely a period during the study where SARS-CoV-2 variants were in circulation; however, strain genotypes for individual cases of breakthrough infections are unknown. As such, a mismatch between the vaccine target protein and the infecting strain may cause a decrease in vaccine effectiveness estimates in the latter part of the study, particularly when there is minimal or no cross protection; however, some degree of vaccine target to circulating strain mismatch is consistent across COVID-19 vaccine studies, based on the rapid mutation and selection pressure of SARS-CoV-2. Finally, a low number of severe disease cases limited the precision of this specific outcome analysis.

Overall, this large real-world assessment of NVX-CoV2373 supplies evidence for its effectiveness in the general population. Data indicate the favorable vaccine effectiveness of NVX-CoV2373 compared with the mRNA-based vaccine, BNT162b2, in preventing any SARS-CoV-2 infection and the potential of an enhanced durable protection from SARS-CoV-2 infection.

## Supporting information

Supplementary Materials for Choe et al

## Author contributions

E.G., S.-A.C., M.V., M.D.R., and Y.J.C. conceptualized the trial and contributed to its design. E.G., S.-A.C., K.K., and E.B. were involved in the extraction and analysis of data. E.G., S.-A.C., K.K., E.B., J.F., M.V., M.D.R., and Y.J.C. were involved in the interpretation of data. E.G., S.-A.C., K.K., E.B., and Y.J.C. verified the data. E.G., S.-A.C., K.K., E.B., J.F., M.V., M.D.R., and Y.J.C. drafted the manuscript. E.G., S.-A.C., and K.K. performed the statistical analyses. S.-A.C., J.F., M.D.R., and Y.J.C. supervised the study. All authors critically reviewed and approved the manuscript.

## Data availability

The data used in this study are from the K-CoV-N cohort, which is not publicly available. Researchers can apply for access to these data through the National Health Insurance Service (NHIS) and Korea Disease Control and Prevention (KDCA) data linkage system. Applications can be submitted via the Health Care Data Linkage (HCDL) website (https://hcdl.mohw.go.kr/) and are subject to review by the relevant authorities.

## Acknowledgments

This study used the Korea Disease Control and Prevention Agency (KDCA) and National Health Insurance Service (NHIS) databases for policy and academic research (KDCA-NHIS-2023-1-494). The conclusions of this study are not related to this institution. Medical writing and editorial support were provided by Kelly M. Fahrbach, PhD, CMPP, and Ebenezer M. Awuah-Yeboah, BS, of Ashfield MedComms (New York, USA), an Inizio company, supported by Novavax, Inc. The authors also thank doctors Mark Thompson and Chengbin Wang for their input on study initiation.

## Appendix A. Supporting information

Supplementary data associated with this article can be found in the online version.

