## Supplementary Materials for Choe et al for "Real-world effectiveness of NVX-CoV2373 and BNT162b2 mRNA COVID-19 vaccination in South Korea"

**Supplement 1**
Matched participant baseline characteristics for the primary series, by vaccine type.

| Variable, n (%) | NVX-CoV2373  (n= 39,762) | BNT162b2  (n=13,380) | aSD^a^ |
| --- | --- | --- | --- |
| Age (year) |  |  |  |
| 12–17 | 568 (1.4) | 269 (2.0) | 0.05 |
| 18–24 | 5526 (13.9) | 1817 (13.6) | 0.01 |
| 25–39 | 15,709 (39.5) | 5831 (43.6) | 0.09 |
| 40–49 | 3781 (9.5) | 1182 (8.8) | 0.02 |
| 50–59 | 2381 (6.0) | 708 (5.3) | 0.02 |
| 60–69 | 3949 (9.9) | 1286 (9.6) | 0.01 |
| 70–79 | 3201 (8.0) | 958 (7.2) | 0.03 |
| ≥80 | 4647 (11.7) | 1329 (9.9) | 0.06 |
| Sex |  |  |  |
| Male | 9093 (22.9) | 2862 (21.4) | 0.03 |
| Female | 30,669 (77.1) | 10,518 (78.6) |  |
| Non-capital area^b^ | 19,512 (49.1) | 6503 (48.6) | 0.01 |
| CCI ≥3 | 2543 (6.4) | 823 (6.2) | 0.01 |
| Disabled | 3670 (9.23) | 1094 (8.18) | 0.04 |
| Employed | 9099 (22.9) | 2715 (20.3) | 0.06 |
| Income level^c^ |  |  |  |
| Medical aid beneficiary | 3008 (8.0) | 856 (6.7) | 0.05 |
| 1Q | 4896 (13.0) | 1555 (12.2) | 0.02 |
| 2Q | 7733 (20.5) | 2655 (20.8) | 0.01 |
| 3Q | 10,629 (28.2) | 3788 (29.7) | 0.04 |
| 4Q | 11,397 (30.3) | 3915 (30.7) | 0.01 |
| Obese (BMI >30 kg/m^2^) | 373 (0.9) | 126 (0.9) | <0.01 |
| Current smoker | 592 (1.5) | 208 (1.6) | <0.01 |
| Prior SARS-CoV-2 infection | 8948 (22.5) | 2919 (21.8) | 0.02 |

^a^aSD values >0.1 indicate a significant imbalance between vaccine groups. ^b^Indication of whether participants were living inside or outside of the Seoul capital area. ^c^Income was categorized as medical aid beneficiary (no/lowest income) and then into quartiles of 10%–40%, >40%–60%, >60%–80%, and >80%–100%.

aSD, absolute standardized difference; BMI, body mass index; CCI, Charleson comorbidity index; Q, quartile.

**Supplement 2**
Any and severe SARS-CoV-2 infection events and adjusted hazard ratios (aHRs) following a homologous primary series (presented by risk window).

|  |  | NVX-CoV2373 | | | BNT162b2 | | |  |
| --- | --- | --- | --- | --- | --- | --- | --- | --- |
| Risk window | SARS-CoV-2 infection | Events, n | Time (1000 person-days) | IR (95% CI)^b^ | Events, n | Time (1000 person-days) | IR (95% CI)^b^ | aHR (95% CI) |
| 30 days | Any | 2122 | 1154 | 1.84 (1.76–1.92) | 925 | 385 | 2.40 (2.25–2.56) | 0.75 (0.69–0.81) |
|  | Severe | 32 | 1192 | 0.02 (0.02–0.03) | 14 | 401 | 0.03 (0.02–0.05) | 0.58 (0.31–1.10) |
| 60 days | Any | 3034 | 2265 | 1.34 (1.29–1.39) | 1321 | 752 | 1.76 (1.66–1.85) | 0.76 (0.71–0.81) |
|  | Severe | 46 | 2383 | 0.02 (0.01–0.02) | 18 | 802 | 0.02 (0.01–0.03) | 0.71 (0.41–1.23) |
| 90 days | Any | 3842 | 3352 | 1.15 (1.11–1.18) | 1633 | 1108 | 1.47 (1.40–1.55) | 0.78 (0.73–0.83) |
|  | Severe | 54 | 3574 | 0.01 (0.01–0.02) | 20 | 1203 | 0.01 (0.01–0.02) | 0.75 (0.45–1.26) |
| 120 days | Any | 4951 | 4412 | 1.12 (1.09–1.15) | 1918 | 1455 | 1.32 (1.26–1.38) | 0.85 (0.81–0.90) |
|  | Severe | 67 | 4464 | 0.01 (0.01–0.02) | 23 | 1603 | 0.01 (0.01–0.02) | 0.81 (0.50–1.31) |
| 150 days | Any | 6290 | 5432 | 1.16 (1.13–1.19) | 2299 | 1793 | 1.28 (1.23–1.34) | 0.91 (0.87–0.96) |
|  | Severe | 84 | 5953 | 0.01 (0.01–0.02) | 26 | 2004 | 0.01 (0.01–0.02) | 0.92 (0.59–1.43) |
| 180 days | Any | 6960 | 6421 | 1.08 (1.06–1.11) | 2523 | 2120 | 1.19 (1.14–1.24) | 0.92 (0.88–0.97) |
|  | Severe | 88 | 7142 | 0.01 (0.01–0.01) | 27 | 2404 | 0.01 (0.01–0.01) | 0.93 (0.60–1.43) |

Risk windows began 14 days post second dose and ran from days 14–44, 14–74, 14–104, 14–134, 14–164, and 14–194. ^b^Calculated per 1000 person-days.

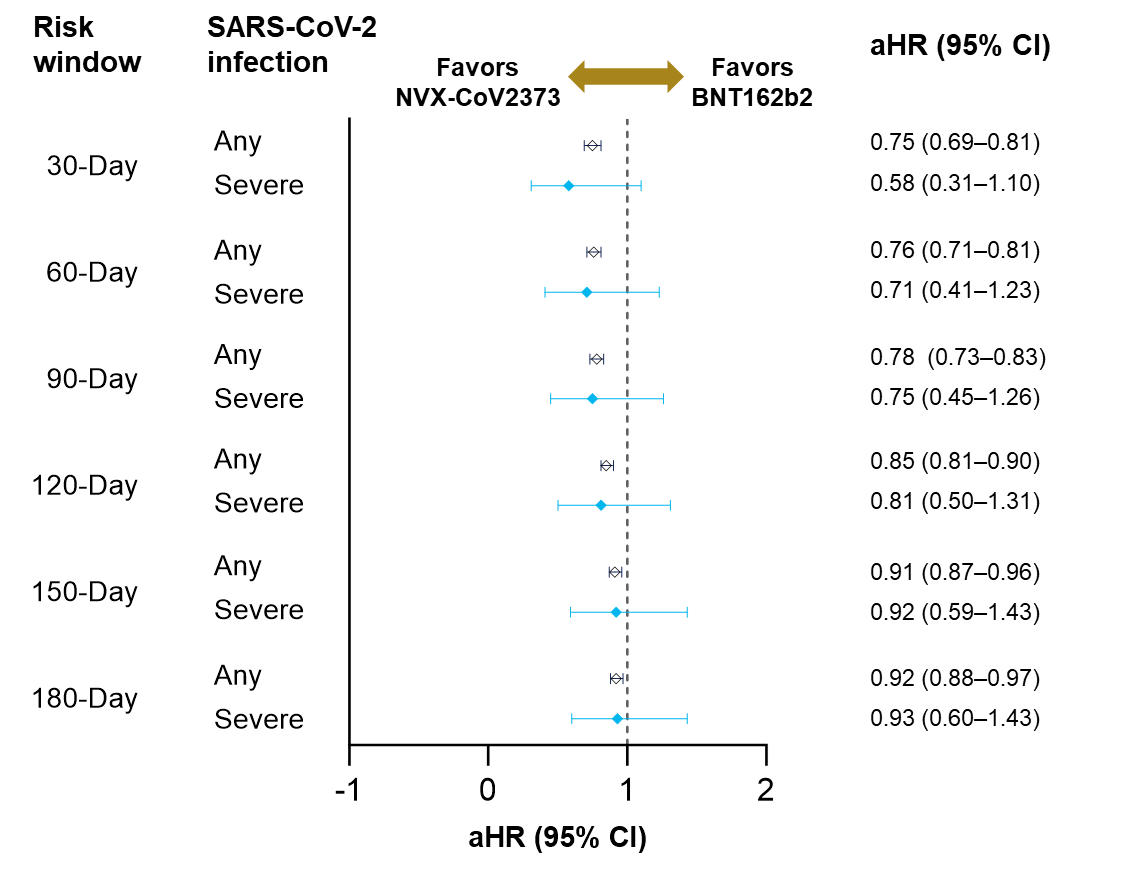

**Supplement 3.** Adjusted hazard ratios (aHRs) of any or severe SARS-CoV-2 infection after a homologous primary series of NVX-CoV-2373 or BNT162b2. Risk windows began 14 days post dose and ran from days 14–44, 14–74, 14–104, 14–134, 14–164, and 14–194.

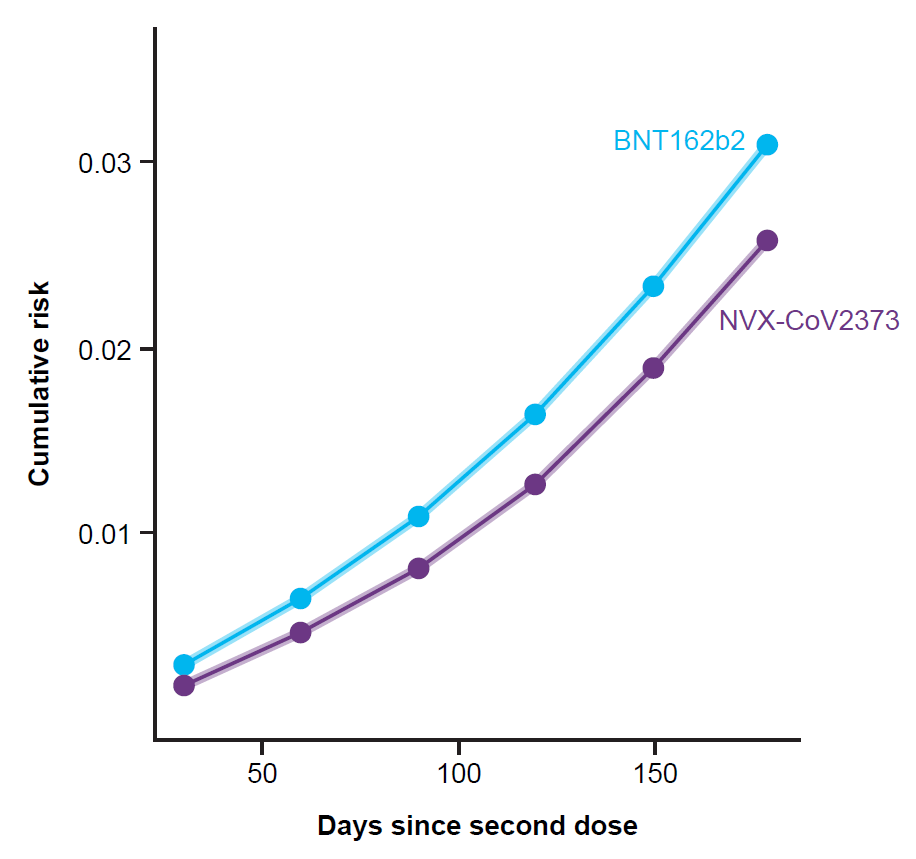

**Supplement 4**. Cumulative risk of any SARS-CoV-2 infection after receipt of a homologous primary series of NVX-CoV2373 or BNT162b2. Shaded areas represent 95% confidence intervals. Risk windows began 14 days post dose and ran from days 14–44, 14–74, 14–104, 14–134, 14–164, and 14–194.

**Supplement 5**
The frequency and adjusted odds ratios (aOR) of all SARS-CoV-2 infections comparing NVX‑CoV2373 and BNT162b2 recipients within 0–5 days post vaccination.

| SARS-CoV-2 infection | NVX-CoV2373 | | BNT162b2 | | *P* | aOR (95% CI) |
| --- | --- | --- | --- | --- | --- | --- |
|  | n | % | n | % |  |  |
| Dose |  |  |  |  |  |  |
| Primary series | 278 | 0.8 | 73 | 0.7 | 0.057 | 1.21 (0.92–1.58) |
| Third dose | 713 | 0.9 | 955 | 0.8 | 0.097 | 1.09 (0.98–1.20) |
| Fourth dose | 1382 | 0.3 | 1167 | 0.2 | <0.001 | 1.16 (1.07–1.25) |
